# Return-to-Play after ACL Reconstruction: A Single-Case Study of Multi-Context Biomechanical Monitoring in Collegiate Basketball

**DOI:** 10.1101/2025.09.23.25336466

**Authors:** Tamar D. Kritzer, Kaylee White, Meaghan Maynard, Anil Palanisamy, Ben Bahrami, Dylan Kobsar

## Abstract

**Introduction:** Anterior cruciate ligament (ACL) injuries represent one of the most common and disruptive conditions in sport, with fewer than two-thirds of athletes return to competitive play. Effective return-to-play (RTP) monitoring requires multidimensional approaches that capture physical, psychological, and sport-specific components rather than reliance of isolated benchmarks.

**Purpose:** This study aimed to longitudinally examine the RTP process of a female varsity basketball athlete following ACL reconstruction, using a framework that integrates physical performance (capacity), biomechanical sport-specific (capability), and psychological (confidence) components relative to pre-injury benchmarks.

**Methods:** Data collection included countermovement jump testing with dual force plates, on-court inertial measurement unit (IMU) monitoring of limb-loading, and psychological questionnaires, analyzed relative to pre-injury and post-surgery baselines using minimal detectable change thresholds.

**Results:** Pre-injury monitoring indicated stable movement profiles with only minor fluctuations. Following ACL reconstruction, jump height recovered within seven weeks of RTP initiation, but notable inter-limb asymmetries persisted in force plate and IMU measures despite high confidence scores.

**Discussion:** Symmetry improved with continued training, yet variability in on-court loading remained even after clinical clearance. These findings highlight the value of integrated, multidimensional monitoring to detect residual deficits that may be overlooked by traditional outcome-based assessments.

**Conclusion:** This study demonstrates that integrating athlete-specific biomechanical, psychological, and sport-specific assessments relative to pre-injury baselines can support RTP decision-making to enhance individualized recovery trajectories in female athletes.

## 1.0 Introduction

Anterior cruciate ligament (ACL) injuries are among the most common and disruptive injuries in sport, accounting for over 400,000 surgeries annually in the U.S. (Devana et al., 2022; Larose et al., 2022; Patterson et al., 2023). Although reconstruction can enable return to sport, recovery affects both physical and psychological well-being, with downstream impacts on satisfaction, knee function, and performance (Patterson et al., 2023). Given that return to play (RTP) is a multi-phase process that extends beyond re-entry to regained performance, and that only 50-65% of athletes in court and field sports return to competitive play (Davies et al., 2020; Jordan & Bishop, 2024), there is a growing need for multi-contextual monitoring that captures readiness across complementary domains rather than single time-point tests alone.

Current RTP guidelines emphasize Capacity, prioritizing strength and power benchmarks. Standard tests, typically administered in clinical or gym settings, include lower-limb strength and hop tests comparing injured and uninjured limbs (Melbourne ACL Protocol). The limb symmetry index (LSI) offers a simple quantification of inter-limb differences but can misrepresent recovery, as both limbs may be affected by injury-related detraining and compensatory strategies often emerge only under sport-specific conditions (Davies et al., 2020; Jordan & Bishop, 2024). To better capture movement quality and task demands, more sport-specific assessments, such as countermovement jump (CMJ) testing on force plates and on-court, game-like evaluations with inertial measurement units (IMUs) may provide deeper insight into neuromuscular strategy and performance (Davies et al., 2020; Jordan & Bishop, 2024; Keogh et al., 2024). Recently, force plate assessments, such as the isometric mid-thigh pull, have been used to evaluate movement quality under higher strength and power demands, helping to identify asymmetries and compensatory strategies (Bishop et al., 2017). Jumping and strength tests conducted with force plates and IMUs have demonstrated reliability for consistent data tracking in real-world training environments, providing objective indicators of movement quality beyond subjective coach observations (Bishop et al., 2017; Keogh et al., 2024). Alongside Capacity, Confidence meaningfully shapes rehabilitation and RTP outcomes. Psychological stressors can compound biomechanical demands and elevate re-injury risk (Keogh et al., 2024). Instruments such as the ACL-Return to Sport after Injury (ACL-RSI) and other questionnaires offer insight into emotional readiness, motivation, and fear of re-injury (Webster & Feller, 2018). Expectations and motivation can influence pain perception and movement confidence, potentially over- or underestimating true readiness and leading to performance setbacks (Faleide et al., 2021; Liu & Noh, 2024).

Despite these advances, a critical gap remains in understanding how Capacity and Confidence evolve over time relative to pre-injury status, and how they align with Capability to perform on court. To address this, we adopt a 3C framework: Confidence, Capacity, and Capability, where Capacity reflects standardized performance (e.g., CMJ metrics) and Capability reflects enacted on-court behavior under real constraints (e.g., IMU-based asymmetry across practice, scrimmage, and games). This framing parallels Glass’s hypothetical, experimental, and enacted function and aligns with unified mobility frameworks (Glass, 1998; Beauchamp et al., 2023). Longitudinal monitoring anchored to pre-injury baselines and minimal detectable change (MDC) can contextualize post-injury shifts, distinguishing typical fluctuations from meaningful deviations that signal risk or recovery (Keogh et al., 2024; Jordan & Bishop, 2024). Accordingly, this single-case study follows a female varsity basketball athlete through the RTP process after ACL reconstruction, leveraging comprehensive pre-injury data to: (1) track changes in movement quality using biomechanical assessments beyond traditional outcome-based approaches; (2) evaluate biomechanical outcomes longitudinally relative to pre-injury baselines using MDC; and (3) examine interrelationships among CMJ force-plate metrics (Capacity), on-court wearable inertial sensor data (Capability), and psychological measures (Confidence) to provide a holistic view of RTP readiness.

## 2.0 Methods

### 2.1 Single-Case Study Design and Phases

This single-case longitudinal study followed a university-aged female varsity basketball player who sustained a contact-related ACL rupture of the left knee during competition. The athlete underwent ACL reconstruction 16 weeks after the date of injury (DOI). Results are organized across three phases: Phase I – Pre-Injury Monitoring (20 weeks pre-DOI), Phase II – Surgery and Rehabilitation (DOI to 32 weeks post-DOI; no biomechanical testing), and Phase III – Return to Play Monitoring (32 weeks post-DOI to the first competitive game of the subsequent season). Study data collection occurred in Phases I and III, using a multi-context framework of Capacity (force-plate CMJ metrics and hop tests), Capability (on-court inertial sensor loading/exposure), and Confidence (psychological self-report). A timeline of injury, surgery, rehabilitation, and data-collection milestones is shown in Figure 1. Detailed protocols of each measure are provided below. This study was reviewed and approved by the university research ethics board.

**Figure 1.**
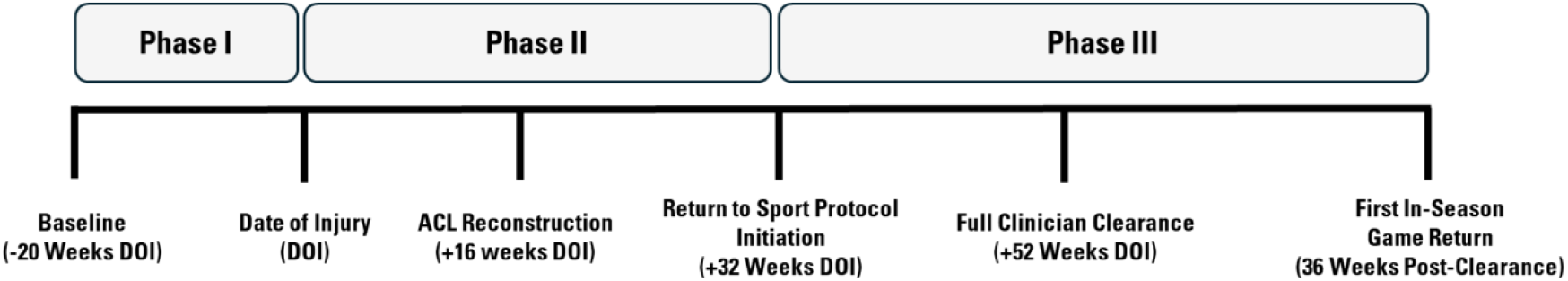
Timeline of injury, surgical intervention, and return-to-play process.

### 2.2 Procedures

#### 2.2.1 Countermovement Jump Assessments

##### Protocol

CMJ assessments were performed twice weekly throughout the assessment period (i.e., Monday and Thursday). On each testing day, the participant completed three jumps, with a minimum of 30-s rest between trials. If a system or individual error occurred, the trial was recollected to ensure reliable and accurate results. The participant was familiar with the CMJ protocol, as it was a routine component of her strength and conditioning program. CMJ testing was conducted at the start of strength and conditioning sessions to ensure the athlete was fully rested and jump performance would not be affected by neuromuscular fatigue. A five-minute standardized warm-up was completed to prepare the neuromuscular system for the task at hand. In each CMJ testing session, research assistants provided instructions to stand still with feet shoulder width apart on the dual force plates and allow for proper establishment of body weight calculation. Then the athlete was provided a verbal cue to “jump as high and quickly as possible” to effectively utilize a stretch–shortening cycle and mimic in-game explosiveness. All jumps were completed with hands placed on hips restricting arm-swing. No instructions were provided for the landing phase aside from ensuring that both feet made contact with the force platform prior to concluding the downward motion of this phase of movement and, subsequently, returning to an upright standing position.

##### Variables of Interest

A dual AMTI force plate system (Hawkins Dynamics, Westbrook, ME, USA) was utilized to measure CMJ force variables bilaterally. A total of 13 movement strategy CMJ variables were obtained for each jump including jump height (m), time to take-off (s), modified reactive strength index (mRSI), peak braking power (W), peak propulsive power (W), left/right peak braking force (N), left/right peak propulsive force (N), left/right average braking RFD (N/s), and left/right peak landing force (N). Movement asymmetry during the CMJ was quantified in percentage by comparing left/right use in four phases: peak braking force, peak propulsive force, average braking RFD, peak landing force.

#### 2.2.2 Inertial Measurement Unit Assessments

##### Protocol

Inertial measurement units (IMUs) were used to gather on-court tri-axial acceleration (g) data of each leg during team practices (IMeasureU Inc., IMU Step, Denver, CO, USA). A single IMU was securely attached above the lateral malleolus on each ankle using elastic straps to ensure stable sensor placement throughout dynamic movement. Data were collected at one to two practices per week across the entire monitoring period of the case study. Following each session, the raw data were wirelessly transferred to a mobile device and uploaded to the IMeasureU cloud platform for automated segmentation and analysis.

##### Variables of Interest

A total of four on-court movement strategy variables were derived. Peak acceleration (g) (Acc_peak_) indicated the max impact of each leg throughout each practice. Acc_peak_ was then further categorized into low (Acc_low_), medium (Acc_med_), and high (Acc_high_) intensities based on number of steps (1-5g, 6-20g, and 21-200g, respectively). Peak impact accelerations were used to calculate between-limb asymmetries, providing insight into limb loading strategies during sport-specific activity.

#### 2.2.3 Psychosocial Clinical Analysis

Psycho-social data was collected and provided by clinicians during the RTP stage (Melbourne ACL Protocol). The ACL-RSI scale was completed six times throughout RTP and the IKDC Subjective Knee Evaluation was completed at the final assessment.

### 2.3 Data Analysis

Data was processed using custom MATLAB code (Matlab R2019a, Mathworks Inc., Natick, MA). The CMJ and on-court data were averaged per day and week to longitudinally analyze changes in movement strategies. If an individual trial had a jump height and weight which deviated by ≥20% and ≥3%, respectively, within a session, the trial was excluded from any further analyses (Keogh et al., 2013). The repeated data collection allowed for tracking of longitudinal trends in movement strategy and asymmetry across the athlete’s rehabilitation and RTP timeline.

### 2.4 Statistical Analysis

Data were statistically analyzed using the custom MATLAB code. To detect statistically significant subject-specific fluctuations from normative baseline patterns, the standardized error of the measurement (SEM) and MDC statistics were computed to identify meaningful levels of change in our biomechanical components across the RTP process (Dvir, 2015; Keogh et al., 2023) (appendix A, Table 1). The SEM was derived from the reliability and standard deviation of repeated measured, while the MDC was calculated from the SEM using the 95% confidence interval (i.e., SEM x 1.96 x √2), reflecting the minimal change beyond expected measurement error. These MDC thresholds were defined using weekly preseason data as normative baseline values from a previous study with the same sample (Keogh et al., 2023). Accordingly, a notable deviation was identified when an athlete’s data exceeded the defined MDC, highlighting a change larger than the expected level of variance for a given component (Dvir, 2015; Keogh et al., 2023).

To explore potential associations between psychosocial clinical assessment outcomes and biomechanical variables during the RTP period, a Spearman’s rank-order correlation was used as a descriptive tool. This analysis aimed to identify patterns of within-individual relationships across time. Correlation coefficients (ρ) were reported for each pairwise comparison to highlight the strength and direction of these associations.

## 3.0 Results

### 3.1 Phase I - Pre-Injury Monitoring

Approximately 20 weeks of pre-injury data were collected during the athlete’s preseason and regular-season period. A standardized data collection schedule was followed as consistently as possible. However, regular fluctuations in varsity team practices, competitions, and travel resulted in some variation in both testing frequency and athlete performance. This variability is expected in an in-season context, where external demands can influence physical readiness and day-to-day performance outcomes.

To aid interpretation, all pre-injury data are presented in Figure 2, with MDC bands delineating typical fluctuations from meaningful change. Variability was most pronounced in asymmetry metrics, consistent with previous literature (Keogh et al., 2024; Jordan & Bishop, 2024). CMJ data showed a consistent slight right-limb bias (∼5%) (Figure 2a-c), while on-court impact loading measured via IMUs was near symmetric (∼1% left-limb bias) (Figure 2d-e). Although the athlete occasionally exceeded MDC thresholds, performance returned to expected ranges, likely reflecting normal responses to training and competition. Overall, no sustained deviations were observed, indicating a relatively stable movement profile throughout the pre-injury phase and providing baselines for post-injury comparisons.

**Figure 2.**
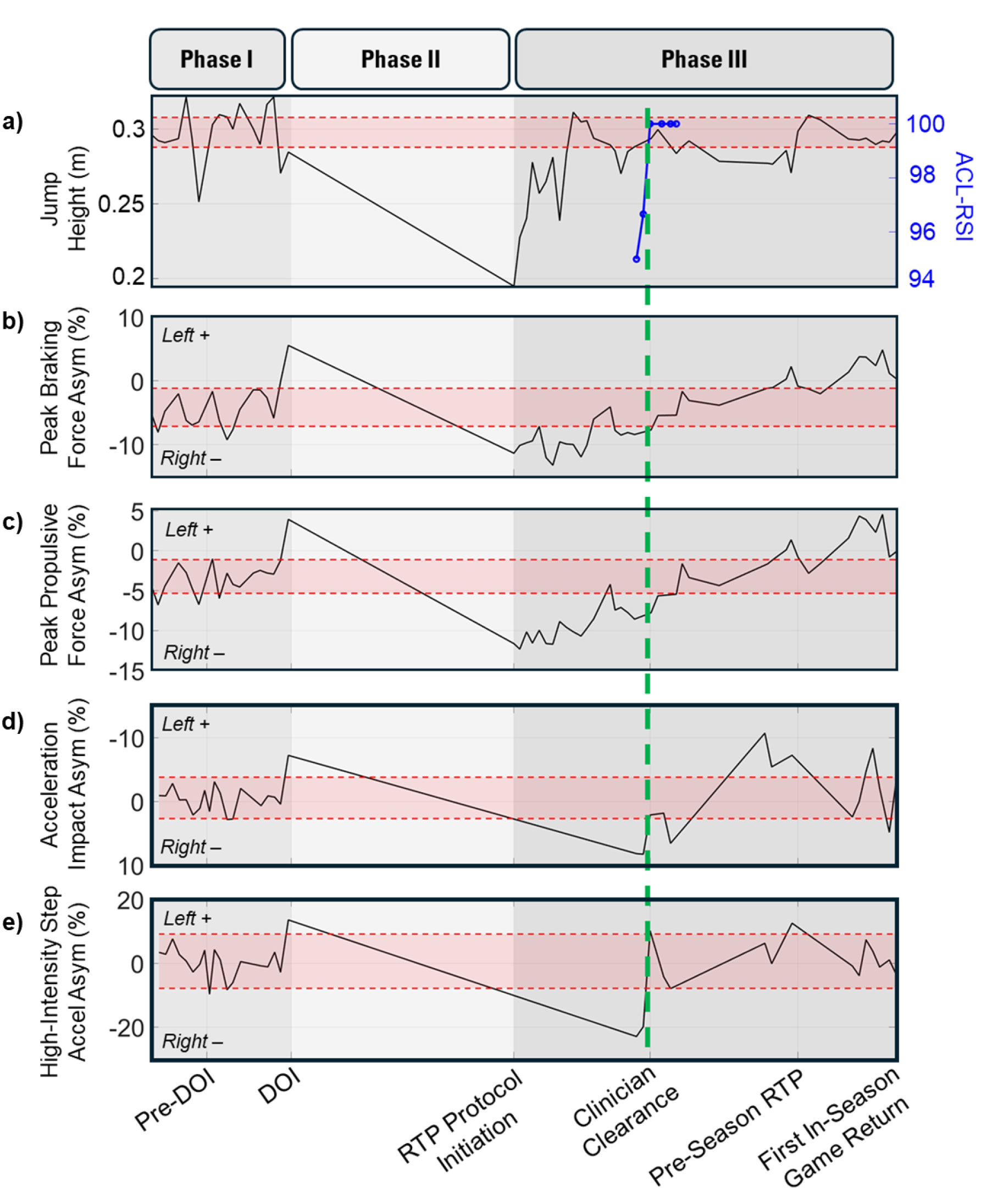
Pre-injury weekly preseason baseline MDC compared to full timeline of injury and RTP recovery. a) CMJ height performance compared to ACL-RSI scores (i.e., clinical knee confidence and functionality patient subjective results. b) CMJ peak braking force asymmetries and c) CMJ peak propulsive force asymmetries demonstrating a gradual return with overcompensations post-clearance. d) IMU acceleration impact asymmetries and e) high-intensity step asymmetries exhibiting fluctuations in on-court movement post-clearance. Date of Injury (DOI): vertical red line, clinical clearance: vertical green line, minimal detectable change bands are shown in red, and ACL-RSI scores are shown by the blue points.

Late in the regular season, the athlete sustained a contact-related ACL injury during a rebound. An opposing player fell onto her left leg, forcing it into hyperextension. The injury was suspected to be a complete ACL tear by the team’s athletic therapist and was subsequently confirmed by an orthopedic surgeon. The athlete’s season ended immediately, and she entered the waiting period for ACL reconstruction surgery. During the 16 weeks preceding surgery, the athlete completed an intensive strength program led by the team’s performance coaches, which played a key role in the rehabilitation process.

### 3.2 Phase II – Surgery and Rehabilitation

The athlete underwent primary ACL reconstruction 16 weeks after the initial injury. A quadriceps graft was used with a lateral meniscal repair using patellar femoral and medial compartment cartilage debridement, with no reported surgical. Post-surgery, she entered a structured rehabilitation program directed by the surgeon and supervised by the team’s athletic therapist, implemented by the strength and conditioning staff, focusing on progressive restoration of mobility, strength, and neuromuscular control, mirroring the Melbourne Protocol.

No biomechanical testing (CMJ or IMU) was performed during this period, as she had not yet advanced to dynamic, sport-specific tasks suitable for these assessments (e.g., running, cutting, and jump-landing). Clearance from this phase required strength and movement assessments, with the affected needing to be within 5% of the unaffected limb in both isolated and multi-joint movements.

Phase II spanned from the date of injury to 32 weeks post-injury, at which point she transitioned to more complex on-court movements and longitudinal biomechanical monitoring resumed in Phase III.

### 3.3. Phase III – Return to Play Monitoring

When functional movement testing resumed during the RTP process, force plate assessments showed a substantial reduction in CMJ height relative to pre-injury levels (Figure 2a). However, jump height improved rapidly, returning to baseline by approximately 7 weeks post-RTP protocol initiation. Despite this recovery in jump height, force-time analyses demonstrated >10% inter-limb asymmetry in braking and propulsive forces, such that the athlete was now relying more on the right (uninjured) limb to achieve pre-injury jump height (Figure 2a-b).

As the athlete moved toward full RTP clearance, she participated in on-court movement sessions emphasizing sport-specific tasks, enabling concurrent IMU monitoring and ACL-RSI reporting. Early IMU sessions showed greater impact-acceleration loading on the uninjured limb (Figure 2d), indicating off-loading of the injured limb and aligning with force plate findings. This was most evident in high-intensity impacts (≥21g; Figure 2e). Notably, this off-loading occurred despite high reported confidence (ACL-RSI = 95). With continued training, inter-limb symmetry improved in both CMJ and IMU measures, coinciding with further gains in confidence as the ACL-RSI reached 100. Although data points were limited, this co-occurrence is illustrated in Figure 3. Finally, to achieve clinical clearance, single-leg hop tests revealed ≥95% symmetry in distance metrics (single hop and triple hop), supporting full RTP clearance (vertical green line, Figure 2).

**Figure 3.**
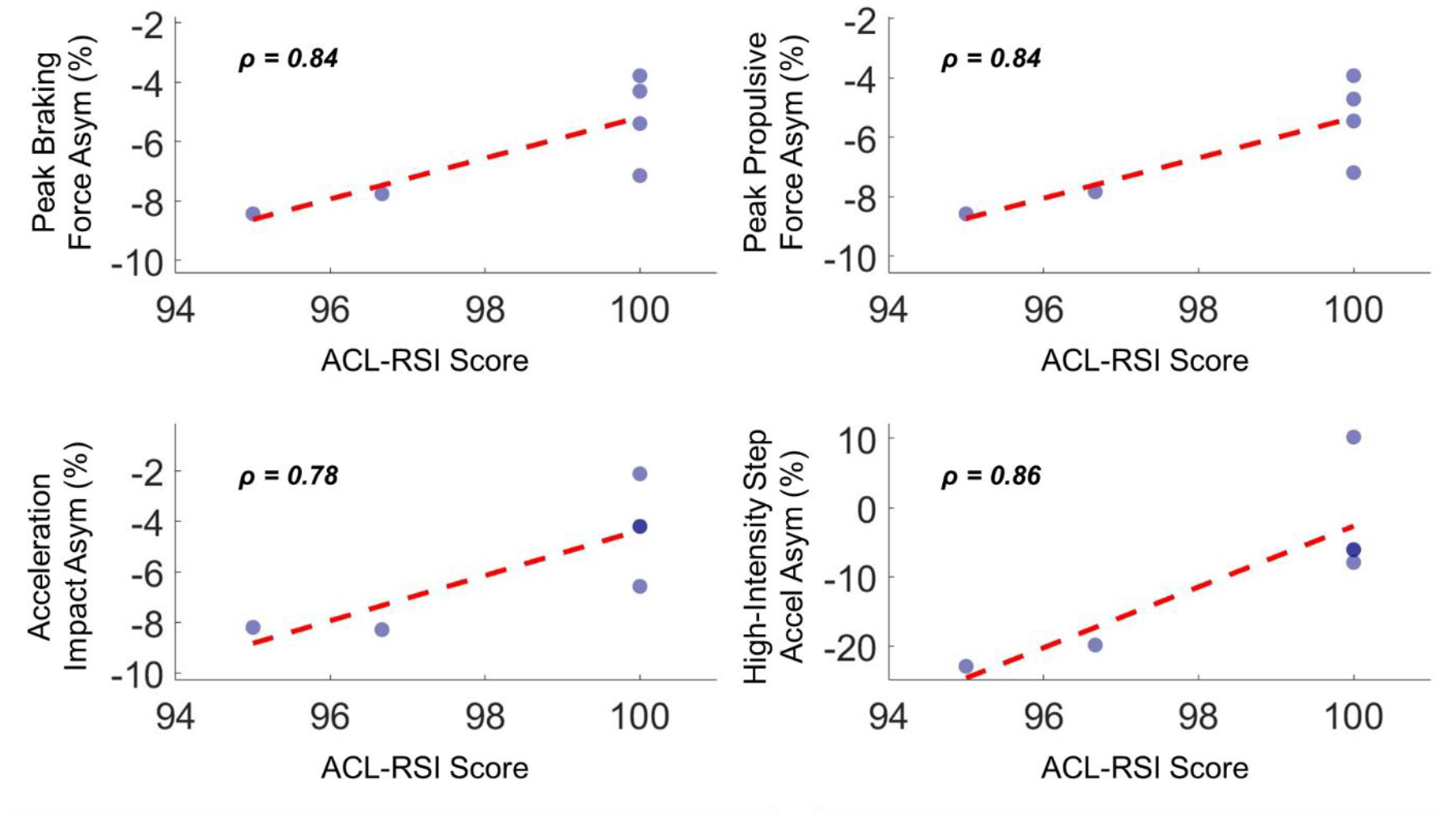
Scatter plots of biomechanical asymmetry measures against ACL-RSI taken near clinician clearance. Spearman correlations presented on each sub-plot.

After clinical clearance, biomechanical markers continued to change. In the weeks leading into preseason, CMJ braking and propulsive asymmetries returned toward pre-injury values with a small residual right bias initially. These asymmetries eventually crossed zero and progressed to a slight left-limb-dominant pattern (∼0-5%) as regular-season play approached (Figure 2b-c). In contrast, IMU-derived on-court loading did not show a consistent directional shift but exhibited variability exceeding pre-injury levels past clinician clearance (Figure 2d-e).

## 4.0 Discussion

The purpose of this single-case study was to longitudinally examine changes in movement quality using biomechanical variables and to provide an individualized, multi-contextual analysis integrating CMJ metrics, on-court IMU loading measures, and psychosocial outcomes. Jump height returned to pre-injury levels early, with hop-test symmetry ≥95% and self-reported knee confidence supporting clinician clearance, despite CMJ and IMU analyses indicating lingering asymmetry favoring loading the uninjured limb. Post-clearance, asymmetry continued to evolve, shifting to a slight left-limb (injured-limb) dominant pattern in CMJ, while on-court loading remained variable. These findings underscore the value of multi-contextual, longitudinal assessments to track alignment across the “3C” framework throughout the RTP process: Confidence (self-report), Capacity (standardized performance tests), and Capability (enacted on-court behavior across practice, scrimmage, and games).

### 4.1 Deficits hidden beyond performance outcomes

RTP protocols that rely solely on traditional performance-based assessments may fail to capture persistent neuromuscular deficits, as they often emphasize outcomes (e.g., jump height or distance) without revealing underlying movement strategies (Davies et al., 2020; Jordan & Bishop, 2024; Keogh et al., 2024). In this single case-study, vertical jump height returned to pre-injury levels early, yet CMJ analyses showed continued overreliance on the uninjured limb (Figure 2a-c). Such discrepancies indicate that this restored performance in jump height did not necessarily reflect normalized movement strategies and may have obscure compensatory patterns that persisted early in recovery (Davies et al., 2020; Jordan & Bishop, 2024; Keogh et al., 2024). Persistent compensation aligns with known post-ACL reconstruction neuromuscular adaptations, such as co-contraction strategies and arthrogenic muscle inhibition, which may protect the graft but produce long-term inter-limb asymmetries and altered mechanics (Nyland et al., 2010; Smeets et al., 2021). Consistent with this, our monitoring showed that the athlete maintained vertical jump height via inter-limb compensation (Figure 2a-c). These findings highlight that restored performance output does not equate to normalized movement strategy and that ongoing asymmetries may remain undetected when relying solely on isolated performance benchmarks in a bilateral vertical jump.

While unilateral assessments may better expose limb-specific deficits (Jordan & Bishop, 2024), their sensitivity may still be limited when they rely only on outcome measures (e.g., hop distance). In this case, the athlete achieved symmetrical single-leg hop test results (>95%) at clinical clearance despite persistent force-time asymmetries in the bilateral CMJ (Davies et al., 2020). Capturing force-plate data during standard single-leg hop tests was not feasible, given the horizontal nature of these tasks typically requires multiple in-ground plates in a laboratory rather than portable, above-ground systems. Consequently, although force-time analysis of single-leg hops would likely be more informative than distance alone, it is often impractical to acquire in applied settings. A pragmatic alternative may be the vertical single-leg hop on a force plate, which enables force-time analysis and symmetry profiling while accentuating unilateral loading demands (Jordan & Bishop, 2024). Future work should evaluate this approach’s sensitivity and reliability alongside bilateral CMJ assessments for RTP monitoring, while acknowledging the potential for substantial inter-limb kinematic variability inherent to single-leg tasks.

Moreover, because compensatory strategies and inter-limb asymmetries are inherently more variable than simple outcome measures, repeated assessments are essential. Outcome metrics such as jump height typically show higher data-to-day reliability than many force-time variables (Keogh et al., 2024), consistent with the MDC values observed here. This stability, while desirable, also underscores jump height’s limited sensitivity to meaningful fluctuations in neuromuscular status, allowing changes to go undetected compared with more detailed force-time measures (McClean et al., 2024). To leverage the greater sensitivity of force-time asymmetries metrics, they should be tracked across multiple time points to distinguish consistent maladaptive trends from single-session outliers that may threaten dynamic stability and performance (Bishop et al., 2013; Bishop et al., 2018).

### 4.2 Individualized vs. standardized symmetry targets

Symmetry targets in rehabilitation may be improved by anchoring benchmarks to an athlete’s natural pre-injury profile rather than enforcing perfect symmetry. Standard clinical protocols often use ≤10% inter-limb asymmetry for RTP, and asymmetries ≥15% have been associated with higher injury incidence (Bishop et al., 2018). However, such cut-points do not account for individual baseline movement strategies, and de-training or strengthening of the contralateral limb after injury can complicate interpretation; apparent symmetry may simply reflect reduced capacity on the previously uninjured side (Jordan & Bishop, 2024). In this case, pre-injury CMJ performance showed a consistent right-leg dominance of ∼3-5%, which was exaggerated early in rehabilitation and later shifted post-clearance toward slight left-leg (injured) reliance (∼0-3%). These observations raise a practical question: should rehabilitation aim for perfect symmetry or a return to the athlete’s characteristic baseline (within MDC limits)? Relying solely on generic thresholds risks overlooking compensatory strategies that diverge from the athlete’s natural profile. Moreover, asymmetries are task- and context-specific (Jordan & Bishop, 2024), where improvements in Capacity during standardized testing may not translate directly to on-court Capability, and vice versa. Accordingly, continuous monitoring of the magnitude and direction of asymmetry across contexts and exposures is essential to guide progression and calibrate RTP decisions.

### 4.3 Variability During RTP

While CMJ asymmetries showed clear trends, on-court IMU monitoring displayed similar directionality with greater session-to-session variability throughout Phase III. These patterns are meaningful as they both reflect the transition of asymmetrical capacity toward symmetry and the challenge of re-establishing consistent on-court performance. Prior to injury, the athlete demonstrated near-symmetrical limb loading amid normal week-to-week fluctuations, likely driven by contextual factors such as session content and exposure (McClean et al., 2024; Jordan et al., 2024; Keogh et al., 2024). Near clinician clearance, the initial on-court loadings shifted away from the reconstructed limb, suggesting an ongoing path towards restoring movement strategies in sport-like conditions, whether consciously or subconsciously (Nyland et al., 2010). This shift aligned with self-reported confidence (Figure 3): as confidence increased, inter-limb symmetry improved on-court, particularly for high intensity impacts (≥21 g; Figure 3) associated with movement like jumps, cuts, and sprints. Once confidence reached its ceiling, directional bias in on-court load was less clear, yet variability remained elevated until closer to the first pre-season game. This heightened variability likely represents a transitional period of re-learning and acclimatizing to higher training loads. Recognizing variability as a meaningful metric, rather than dismissing it as noise, and tracking its magnitude and trajectory longitudinally may provide further insight into neuromuscular control and adaptability, an underexamined aspect of RTP assessment.

### 4.4 Integrating Confidence, Capacity, and Capability for Comprehensive RTP

Alignment across Confidence, Capacity, and Capability is central to understanding RTP. Traditional protocols emphasize Capacity (performance under standardized tests) and, increasingly, Confidence (self-reported readiness), yet these domains do not always translate to on-court Capability (enacted function in sport-specific environments). In practice, an experienced coach or trainer may synthesize these signals, but gaps can arise when information is siloed or when providers have differing vantage points. For example, strength and conditioning staff may detect Capacity limitations that are not evident to coaching staff focused on practice performance, whereas coaches may observe on-court Capability that outpaces underlying neuromuscular Capacity. Confidence is additionally hard to appraise, and athletes may over- or under-report it; psychological readiness has been linked to functional performance and mechanics, with lower scores associated with asymmetrical loading and altered knee moments (Ardern et al., 2016; Peebles et al., 2021).

A multi-context, longitudinal approach, explicitly integrating Confidence, Capacity, and Capability, can reduce these blind spots, paralleling comprehensive function frameworks in mobility research (Glass, 1998; Beauchamp et al., 2023). In our case, ACL-RSI reached a ceiling while CMJ symmetry and on-court loading were still realigning, illustrating temporary discordance across domains and the potential for motivated athletes to modify strategies during single-session tests. Routine alignment checks across the three domains (e.g., ACL-RSI within target; CMJ/hop symmetry within MDC of pre-injury; on-court IMU loading/asymmetry within pre-injury variability at comparable exposures) could flag when progression is outpacing underlying readiness. Operationally, integrated support teams might require agreement in at least two domains before clearance and then continue targeted monitoring of the third. Such integration supports individualized rehabilitation, more efficient movement strategies, and, ultimately, mitigation of re-injury or contralateral risk.

## Conclusion

Given the higher incidence of ACL injuries and poorer post-surgical outcomes observed in female athletes, driven by sex-specific anatomical, neuromuscular, and strength-related factors, it is critical that RTP protocols adopt an individualized and holistic approach to rehabilitation (Agel et al., 2007; Waters, 2012; Tramer et al., 2020; Figueroa et al., 2024). This case study illustrates how integrating longitudinal, athlete-specific biomechanical assessments with psychological and performance variables can enhance clinical decision-making and support safer, more effective RTP trajectories for female athletes (Keogh et al., 2023). Incorporating pre-injury baseline data and monitoring movement strategy adaptations over time allows practitioners to identify compensations and potential reinjury risks that standardized timelines may overlook. Future research and clinical practice should prioritize tailored, multidimensional RTP frameworks that address the unique recovery profiles of female athletes to reduce reinjury risk and optimize long-term outcomes.

## Data Availability

All data produced in the present study are available upon reasonable request to the authors.

## Appendix A

**Table 1.** Minimal Detectable Change Statistical Results.

|  | **ICC** | **SEM** | **MDC** |
| --- | --- | --- | --- |
| Jump Height (m) | 0.99 | 0.0051 | 0.01 |
| Time To Takeoff (sec) | 0.97 | 0.02 | 0.06 |
| mRSI | 0.97 | 0.0093 | 0.03 |
| Peak Braking Power (W) | 0.98 | 39.86 | 110.5 |
| Peak Braking Force Asymmetry Index (N%) | 0.98 | 1.08 | 2.98 |
| Left Force at Peak Braking Force (N) | 0.99 | 15.16 | 42.03 |
| Right Force at Peak Braking Force (N) | 0.98 | 19.28 | 53.44 |
| Peak Propulsive Power (W) | 0.99 | 34.33 | 95.16 |
| Peak Propulsive Force Asymmetry Index (N%) | 0.97 | 0.76 | 2.11 |
| Left Force at Peak Propulsive Force (N) | 0.99 | 10.56 | 29.28 |
| Right Force at Peak Propulsive Force (N) | 0.98 | 13.31 | 36.9 |
| Average Braking RFD Asymmetry Index (N/s) | 0.97 | 2.39 | 6.62 |
| Average Left Braking RFD (N/s) | 0.95 | 177.61 | 492.31 |
| Average Right Braking RFD (N/s) | 0.94 | 195.17 | 540.99 |
| Peak Landing Force Asymmetry Index (N%) | 0.92 | 4.41 | 12.21 |
| Left Force at Peak Landing Force (N) | 0.97 | 53.89 | 149.39 |
| Right Force at Peak Landing Force (N) | 0.84 | 75.19 | 208.41 |
| Impact Load Total ($Acc_{peak}$) (g) | 0.93 | 8,385 | 16,435 |
| Impact Asymmetry (g%) | 0.89 | 1.66 | 1.93 |
| Step Count Difference – Low Intensity (1-5g) Asymmetry Index (%) | 0.83 | 0.99 | 3.24 |
| Step Count Difference – Medium Intensity (6-20g) Asymmetry Index (%) | 0.95 | 0.87 | 1.70 |
| Step Count Difference – High Intensity (21-200g) Asymmetry Index (%) | 0.84 | 4.34 | 8.51 |

Abbreviations: ICC = Intraclass Correlation Coefficient; SEM = Standard Error of Measurement; MDC = Minimal Detectable Change.

